# Machine Learning Prediction of Blood Pressure Control in Patients With Hypertension and Heart Failure Using Longitudinal Clinical Data

**DOI:** 10.64898/2025.12.25.25343007

**Authors:** Haoxin Chen, Jiancheng Ye

**Affiliations:** Weill Cornell Medicine, Cornell University, New York, NY, United States; Northwestern University, Chicago, IL, United States

**Keywords:** Hypertension, blood pressure, machine learning, electronic health records, heart failure, predictive modeling

## Abstract

**Objective:** To develop and validate machine learning models for predicting Blood Pressure (BP) control status using demographic characteristics and longitudinal BP history.

**Methods:** This retrospective cohort study analyzed deidentified data from a multi-site primary care quality improvement program for hypertension management. Participants included adults with diagnosed hypertension (N=23,002) or heart failure (N=1,137) who had at least 2 clinical visits. The primary outcome was BP control status, defined as systolic BP less than 140 mm Hg and diastolic BP less than 90 mm Hg. Five machine learning algorithms (logistic regression, decision tree, Random Forest (RF), support vector machine, and extreme gradient boosting) were compared using accuracy, precision, recall, F1-score, and area under the receiver operating characteristic curve (AUROC). Feature importance was assessed using Shapley Additive Explanations (SHAP) values.

**Results:** Among 23,002 hypertensive patients (mean [SD] age, 65.25 [13.95] years; 13,015 [56.58%] female), the RF model achieved the highest performance with an AUROC of 0.88 (95% CI, 0.86-0.90) using BP history features alone and 0.87 (95% CI, 0.85-0.89) with combined features. BP history substantially outperformed demographic factors (AUROC, 0.60; 95% CI, 0.58-0.62). Mean systolic BP, maximum systolic BP, and maximum diastolic BP were the most influential predictors. In the heart failure cohort (N=1,137; mean [SD] age, 75.15 [15.05] years; 579 [50.92%] female), the RF model achieved an AUROC of 0.93 (95% CI, 0.90-0.96) with combined features. The model demonstrated accuracy of 0.77 (95% CI, 0.76-0.78), precision of 0.78 (95% CI, 0.76-0.79), recall of 0.73 (95% CI, 0.71-0.75), and F1-score of 0.75 (95% CI, 0.74-0.77) for the hypertension cohort.

**Conclusions:** Machine learning models incorporating longitudinal BP history effectively predicted hypertension control status, with BP variability metrics showing substantially greater predictive value than demographic characteristics. These findings suggest that systematic incorporation of historical BP patterns into clinical decision-support systems may enhance personalized hypertension management.

## INTRODUCTION

Hypertension represents a critical global public health challenge. The growing burden of this condition has significant implications for public health, as uncontrolled hypertension is a leading risk factor for severe cardiovascular events. According to the World Health Organization (WHO), an estimated 1.4 billion adults aged between 30 and 79 worldwide have hypertension in 2024, but only 23 % of them have it under control [1]. This alarming control gap has profound implications, as uncontrolled hypertension serves as a leading modifiable risk factor for cardiovascular morbidity and mortality, including stroke, myocardial infarction, heart failure, and chronic kidney disease.[2] Optimal hypertension management necessitates regular clinical monitoring and iterative therapeutic adjustment, rendering healthcare visit patterns and longitudinal blood pressure (BP) trends essential components of care quality assessment and outcome prediction.[3]

Heart failure constitutes another major cardiovascular disease burden with significant overlap with hypertension, affecting approximately 26 million individuals globally.[4] In the United States alone, nearly 6.7 million adults aged 20 years or older currently live with heart failure, a condition that contributed to approximately 14.6% of all-cause mortality in recent epidemiological surveys. The economic burden is substantial, with an estimated $30.7 billion in annual healthcare expenditures attributed to heart failure-related services, medications, and productivity losses.[5] Effective heart failure management requires comprehensive, multifaceted approaches encompassing pharmacological therapy with guideline-directed medical therapy, dietary modification including sodium restriction, structured physical activity programs, and in selected cases, advanced interventions such as cardiac transplantation or mechanical circulatory support.[6] Success in heart failure management fundamentally depends on patients’ consistent self-monitoring of symptoms and sustained collaboration with multidisciplinary healthcare teams.

Recent methodological advances in machine learning have demonstrated considerable potential for enhancing predictive modeling capabilities across diverse clinical settings, addressing inherent limitations of traditional statistical approaches in forecasting complex patient outcomes. Zhang et al. incorporated electronic health record (EHR) audit log data to improve hospital-wide discharge predictions, demonstrating that models leveraging granular user-EHR interaction patterns alongside conventional demographic and clinical features achieved superior performance with an AUROC of 0.921, compared to models lacking EHR interaction data (AUROC: 0.862).[7] This work highlighted the substantial predictive potential embedded in routinely collected but often underutilized audit logs. Similarly, Yan et al. developed and rigorously validated a machine learning model to forecast transitions between acute brain dysfunction (ABD) and ABD-free states in intensive care unit (ICU) populations, achieving an AUROC of 0.824 and emphasizing the utility of machine learning in supporting critical resource allocation decisions.[8] Li et al. employed the Shapley Additive Explanations (SHAP) methodology to elucidate an extreme gradient boosting (XGBoost) model, revealing key prognostic factors including blood urea nitrogen as critical determinants of ICU mortality in heart failure patients. Their XGBoost model outperformed alternative machine learning approaches with an AUROC of 0.824, demonstrating potential for more accurate risk stratification and individualized treatment planning in intensive care settings.[9] Likewise, in ICU patients with acute kidney injury (AKI), Liu et al. found that XGBoost demonstrated the highest predictive performance with an AUROC of 0.796, with the model’s superior discrimination of high-risk patients emphasizing its potential for facilitating early intervention and improving patient outcomes.[10] In the context of acute ischemic stroke (AIS), Wang et al. developed machine learning models to predict neurological prognosis, with their Random Forest (RF) model achieving the highest AUROC of 0.908 and identifying key biomarkers such as neuron-specific enolase (NSE) and C-reactive protein (CRP) as significant predictors of poor outcomes.[11]

Advancements in medical AI for hypertension management have also been explored in recent literature. Ma et al. investigated predictors of hypertension control in Chinese populations, identifying elevated body mass index and family history of hypertension as factors associated with poor BP control, while married status appeared to confer protective effects; a support vector machine (SVM) model optimized in their study achieved a prediction accuracy of 80.9%, demonstrating the practical utility of machine learning in hypertension management contexts.[12] Kohjitani et al. comprehensively reviewed the potential of artificial intelligence models for individualized hypertension treatment, with particular emphasis on time-series BP data. They highlighted that the transition from snapshot measurements to continuous time-series data, facilitated by increasing digitalization of daily BP monitoring, enables more accurate characterization of BP variability. Contemporary AI models have demonstrated promise in forecasting BP fluctuations for periods extending up to four weeks, with the growing application of explainable AI techniques enhancing model interpretability and clinical acceptability. These methodological advances are crucial for developing personalized intervention plans. However, the authors identified persistent challenges in validating AI-driven interventions in real-world settings and integrating diverse AI methodologies, necessitating continued interdisciplinary collaboration among AI specialists, clinicians, and healthcare system administrators.[13]

These studies collectively demonstrate the substantial utility of machine learning in developing reliable prognostic tools capable of supporting clinical decision-making and enabling personalized treatment strategies. They also underscore the critical importance of improving predictive model accuracy while simultaneously offering interpretability through methods like SHAP, making these sophisticated models more feasible for clinical adoption and integration into existing healthcare workflows. However, despite considerable advancements in predictive modeling for cardiovascular diseases, a significant gap persists in research systematically exploring how patients’ longitudinal healthcare visit records and BP history quantitatively influence hypertension and heart failure outcomes. While regular clinical visits are intuitively recognized as crucial in common clinical practice, understanding how patients’ comprehensive healthcare history, particularly the temporal patterns and variability in their BP measurements, affects long-term disease management has not been thoroughly investigated using rigorous quantitative methods. Addressing this knowledge gap could yield valuable insights into how patient history influences treatment effectiveness, risk stratification, and ultimately patient outcomes.

This study aims to bridge this critical gap by systematically examining the impact of both demographic characteristics and longitudinal BP history on hypertension and heart failure outcomes. By leveraging machine learning techniques to analyze complex patient data patterns, we seek to uncover previously unrecognized relationships and factors that could meaningfully enhance treatment strategies, improve risk stratification accuracy, and optimize overall patient outcomes in cardiovascular disease management.

## MATERIALS AND METHODS

### Data source

This study utilized deidentified data from a quality improvement program (EvidenceNOW) implemented across multiple primary care practices focused on enhancing hypertension management. Diagnosis data included diagnosis dates, diagnostic terminology, and standardized codes from the International Classification of Diseases (ICD) and Systematized Nomenclature of Medicine Clinical Terms (SNOMED CT), enabling identification of hypertension and related comorbidities including type 2 diabetes mellitus, coronary artery disease, and prior myocardial infarction.

Demographics contained patient-specific sociodemographic information including age at diagnosis, biological sex, self-reported race and ethnicity, marital status, preferred language for healthcare communication, and geographic data including patient residential zip codes. Vital signs provided measurements of key physiological indicators recorded across multiple clinical encounters, including systolic and diastolic BP readings, heart rate, body mass index (BMI), and body weight. Medication data detailed pharmacological interventions prescribed during each clinical visit, including medication name, standardized medication codes, prescribed dosing frequency, and start and end dates of prescriptions.

### Patient selection and data processing

We extracted a comprehensive BP history data for patients with hypertension who had completed at least two clinical visits, incorporating derived variables including mean systolic and diastolic BP across all visits, maximum and minimum BP values, visit frequency (total number of encounters), temporal patterns of care (duration between consecutive visits, days since most recent visit), and visit interval variability. These longitudinal features were integrated with demographic measures to create a complete analytical dataset. We applied an analogous systematic process to identify and characterize the heart failure cohort using established heart failure diagnostic codes, generating a parallel comprehensive dataset for patients with at least two documented clinical visits for subsequent comparative analyses.

### Feature engineering and model development

To systematically investigate factors influencing hypertension and heart failure outcomes while specifically assessing the independent and synergistic contributions of BP history, we evaluated three distinct feature groups: (1) BP history variables exclusively, encompassing longitudinal BP measurements and visit pattern characteristics; (2) demographic factors exclusively, including age, sex, race, ethnicity, preferred language, and geographic location; and (3) a comprehensive combination incorporating both BP history and demographic factors. This structured approach enabled systematic assessment of each feature domain’s relative predictive contribution.

The complete dataset was randomly partitioned into training (80%) and testing (20%) subsets using stratified sampling to maintain outcome class distributions. The training cohort was utilized for model construction and hyperparameter optimization through cross-validation, while the held-out testing set was reserved exclusively for unbiased validation of final model predictive performance. To perform BP control status prediction, we systematically compared five established machine learning algorithms representing diverse modeling paradigms: Logistic Regression (LR), Decision Tree (DT), Random Forest (RF), Support Vector Machine (SVM), and Extreme Gradient Boosting (XGBoost). Each algorithm was selected to represent distinct approaches to classification problems:

Logistic Regression (LR) is a widely utilized parametric model for estimating the probability of binary outcomes and has become a standard approach for clinical classification problems. For multivariate analysis, LR enables modeling of binary dependent variables such as disease presence or absence by estimating regression coefficients for each predictor variable while adjusting for other covariates. The model provides interpretable odds ratios representing the relative risk or odds of an outcome associated with each predictor, making it particularly valuable for clinical applications requiring transparent decision-making. Decision Tree (DT) is a non-parametric model that classifies observations by recursively partitioning data into increasingly homogeneous subsets based on feature values, ultimately generating a tree-like hierarchical structure of decision rules. Decision trees are particularly useful for developing straightforward, highly interpretable models where each branch represents a clinically meaningful decision point, such as determining treatment pathways based on patient symptoms, vital signs, or laboratory results. The final classification decision occurs at terminal leaf nodes, which can predict outcomes such as disease progression, treatment response, or complication risk. RF is an ensemble learning method that constructs multiple decision trees during training and aggregates their outputs through majority voting or averaging to improve prediction accuracy and substantially reduce overfitting. RF algorithms can effectively handle large, complex datasets such as comprehensive electronic health records by capturing intricate interactions among multiple predictor variables. This approach provides robust predictions for diverse clinical outcomes including patient survival rates, hospital readmission risk, and treatment response, offering clinicians a powerful yet relatively interpretable tool for clinical decision support. SVM is a sophisticated classification algorithm that identifies the optimal hyperplane maximally separating classes in high-dimensional feature space. In clinical applications, SVM’s ability to handle high-dimensional data and model complex non-linear relationships through kernel functions makes it an effective tool for predicting outcomes in challenging cases, such as identifying patients at elevated risk for postoperative complications or adverse drug reactions. The kernel trick enables SVM to implicitly transform features into higher-dimensional spaces where linear separation becomes possible. Extreme Gradient Boosting (XGBoost) is an advanced ensemble technique that sequentially constructs decision trees in an additive manner, with each subsequent tree attempting to correct errors made by the ensemble of previous trees. The algorithm incorporates sophisticated regularization techniques to prevent overfitting, can inherently handle missing data through learned optimal default directions, and efficiently models complex interactions among multiple predictors. In clinical practice, XGBoost has demonstrated exceptional efficiency and accuracy in predicting outcomes from large, heterogeneous datasets, such as forecasting patient outcomes following surgical procedures, predicting disease recurrence probability, or identifying patients likely to benefit from specific therapeutic interventions.

All models were implemented using scikit-learn (version 1.0.2) and XGBoost (version 1.5.0) libraries in Python 3.9. Hyperparameters for each algorithm were optimized using 5-fold cross-validation with grid search on the training set to identify configurations maximizing AUROC. Models were trained on the complete training set using optimized hyperparameters before final evaluation on the held-out test set.

### Model evaluation

Each model’s predictive performance was comprehensively assessed using multiple complementary classification metrics, providing a multifaceted evaluation of model quality. Accuracy represents the proportion of correctly classified observations among all observations, providing an overall measure of classification correctness. Precision quantifies the proportion of true positive predictions among all positive predictions made by the model, reflecting the model’s positive predictive value and indicating the probability that a patient predicted to have uncontrolled BP actually has uncontrolled BP. Recall (sensitivity) measures the proportion of actual positive cases that were correctly identified by the model, representing the model’s ability to detect all true positive cases and indicating what proportion of patients with truly uncontrolled BP are correctly identified. The F1-score is computed as the harmonic mean of precision and recall, providing a single balanced metric that accounts for both false positives and false negatives, particularly valuable when class distributions are imbalanced. The AUROC curve illustrates the fundamental trade-off between sensitivity (recall) and specificity across all possible classification thresholds, with values ranging from 0.5 (no discrimination) to 1.0 (perfect discrimination).

For each performance metric, 95% confidence intervals were calculated using bootstrap resampling with 1,000 iterations to quantify estimation uncertainty. For visualization and comparison purposes, AUROC curves for all five machine learning models within each feature group were plotted together, enabling direct visual assessment of relative discriminative performance across modeling approaches.

### Model interpretation using SHAP

To interpret the best-performing machine learning models and elucidate which features most substantially influenced predictions, we applied the Shapley Additive Explanations (SHAP) framework. SHAP values, grounded in cooperative game theory, quantify each feature’s contribution to individual predictions by computing the marginal contribution of each feature across all possible feature coalitions. This approach provides both local (instance-level) and global (population-level) interpretability while satisfying desirable theoretical properties including local accuracy, missingness, and consistency. Based on model performance evaluated by AUROC, we selected the best-performing model and calculated SHAP values for each feature across all observations in the test set. We computed mean absolute SHAP values for each feature, representing the average magnitude of each feature’s impact on predictions across the entire dataset. Features were then ranked according to their mean absolute SHAP values to identify the most influential risk factors and predictors of BP control status. Additionally, we generated SHAP summary plots illustrating the distribution of SHAP values for each feature, with color indicating feature magnitude (high values in red, low values in blue) and horizontal position indicating the direction and magnitude of impact on the predicted outcome.

### Statistical analysis

Continuous variables are presented as mean ± standard deviation (SD), while categorical variables are expressed as frequencies with corresponding percentages. Between-group comparisons for continuous variables were performed using independent samples t-tests (for normally distributed variables) or Mann-Whitney U tests (for non-normally distributed variables), as appropriate based on distributional assessments. Categorical variables were compared using chi-square tests or Fisher’s exact tests when expected cell counts were less than 5. Statistical significance was defined using a two-tailed alpha level of 0.05, with 95% confidence intervals (CIs) calculated for all point estimates. All statistical analyses, including descriptive statistics, demographic and clinical characteristic comparisons, and machine learning model development, were conducted using Python 3.9 with standard scientific computing packages including pandas (version 1.3.3), numpy (version 1.21.2), scipy (version 1.7.1), scikit-learn (version 1.0.2), XGBoost (version 1.5.0), and SHAP (version 0.40.0).

### Study Approval

This study utilized the dataset from a quality improvement program (EvidenceNOW). All the data were de-identified and the study was approved by Northwestern University’s Institutional Review Board.

## RESULTS

### Hypertension cohort

We identified 26,919 patients with documented hypertension diagnoses in the dataset. Of these, 23,002 patients (85.5%) had completed at least two clinical visits and were included in the final analytical cohort. The cohort composition included 13,015 females (56.58%), 9,986 males (43.41%), and 1 patient (0.01%) with undifferentiated or missing sex documentation. The mean age of female patients was 65.80 ± 14.08 years, significantly older than male patients at 64.53 ± 13.74 years (P<0.001), reflecting established epidemiological patterns of hypertension prevalence and healthcare utilization by sex. Substantial and statistically significant demographic differences were observed between male and female patients across multiple characteristics (**Table 1**). Regarding ethnicity, a higher proportion of female patients (53.92%) identified as not Hispanic or Latino compared to male patients (49.97%), while 39.43% of males and 35.54% of females identified as Hispanic or Latino (P<0.001). Racial distribution also differed significantly between sexes (P<0.001). Among patients who reported specific racial categories (rather than “Other”), a higher proportion of females identified as Black or African American (3.31% vs 3.02%), while higher proportions of males identified as Asian (3.24% vs 2.52%) or White (12.66% vs 9.70%). Preferred language for healthcare communication also varied significantly by sex (P<0.001). English was the most commonly spoken language in both groups, reported by 51.88% of females and 46.88% of males. Spanish was the second most common language, spoken by 25.63% of females and 26.67% of males.

**Table 1.**
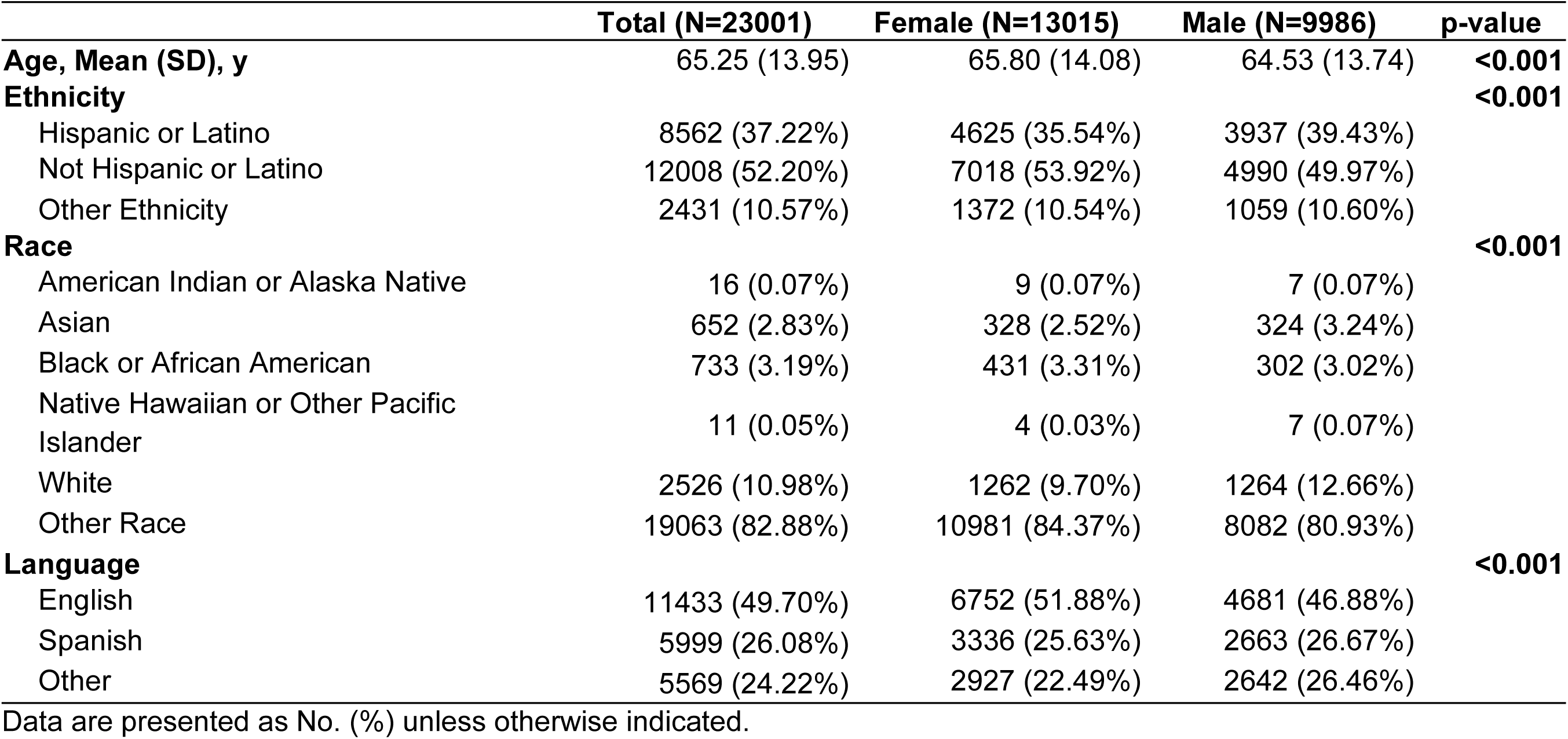
Demographic differences by sex (hypertension cohort)

Regarding BP and clinical visit characteristics (**Supplemental Table 1**), the cohort demonstrated substantial variability in BP control. The mean of each patient’s average systolic BP across all visits was 135.76 mmHg, with a mean of average diastolic BP of 81.86 mmHg. The mean of maximum systolic BP recorded for each patient was 155.69 mmHg, while the mean of minimum diastolic BP was 70.29 mmHg, illustrating considerable within-patient BP variability over time. Patients completed an average of 10.61 visits during the observation period, with a mean of 130.11 days since their most recent visit at the time of data extraction. The median duration between consecutive visits averaged 65.50 days, with substantial variability in inter-visit intervals (SD = 71.25 days) reflecting diverse patterns of healthcare engagement and follow-up intensity.

### Model performance: hypertension cohort BP history features only (group 1)

When models were trained exclusively using BP history and visit pattern features, RF and SVM achieved the highest accuracy and precision, both reaching 0.78 (**Table 2**). Logistic Regression (LR) demonstrated the highest recall at 0.76, indicating superior sensitivity in identifying patients with uncontrolled BP. Decision Tree (DT) showed notably inferior performance compared to other algorithms, likely due to overfitting to training data. Four of the five models (LR, RF, SVM, and XGBoost) achieved identical F1-scores of 0.76, reflecting a balanced trade-off between precision and recall. The AUROC curves for these predictive models (**Figure 1A**) revealed that RF achieved the best discriminative performance among all algorithms with an AUROC of 0.88, demonstrating strong ability to distinguish between controlled and uncontrolled hypertension using BP history alone.

**Fig 1A.**
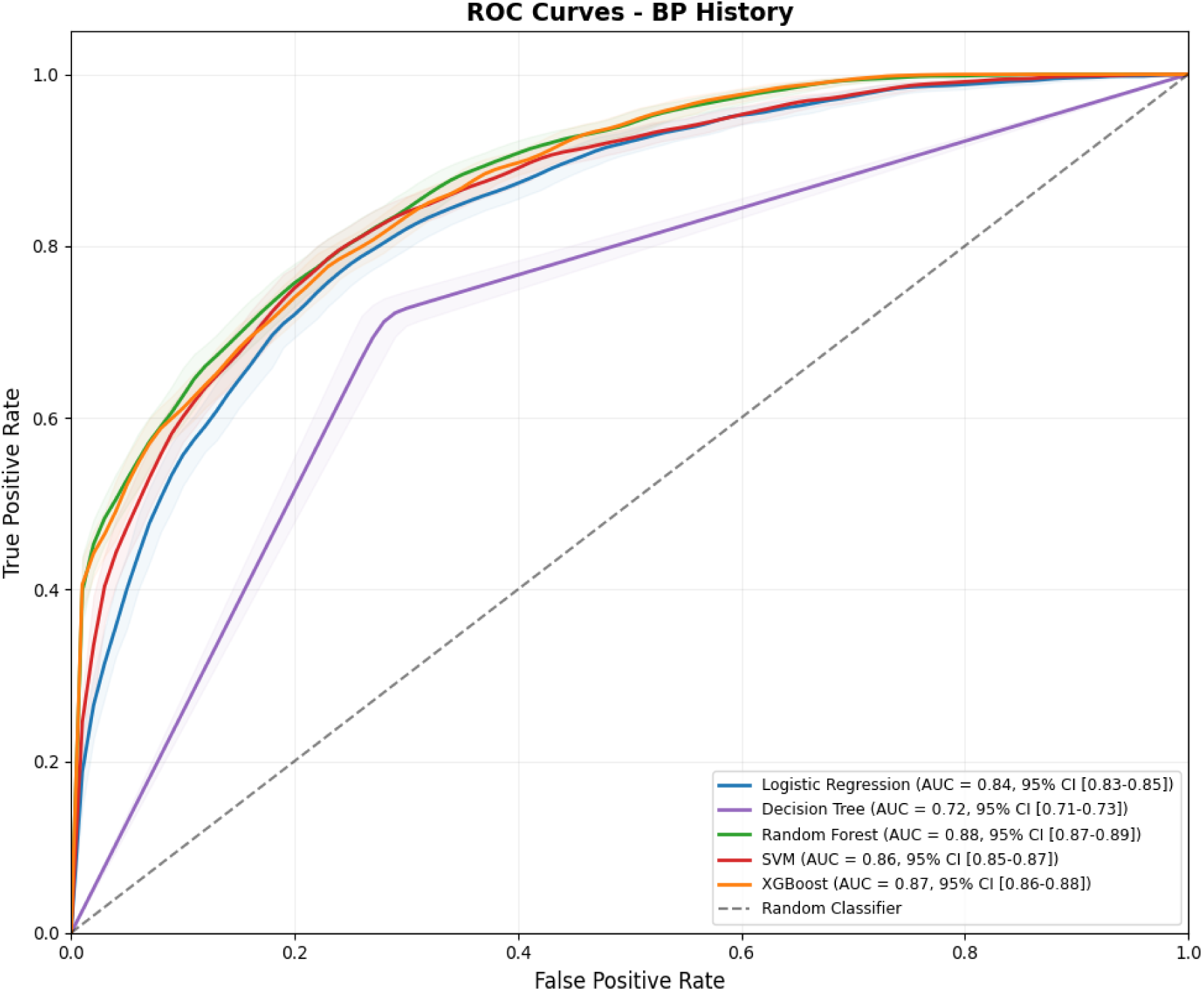
AUROC among five models of BP history (hypertension cohort)

**Table 2.** Models’ performance of three feature groups (hypertension cohort)

| Feature group | Model | Accuracy | Precision | Recall | F1-score |
| --- | --- | --- | --- | --- | --- |
| **BP history** | LR | 0.76 (0.75 - 0.78) | 0.75 (0.73 - 0.77) | 0.76 (0.75 - 0.78) | 0.76 (0.74 - 0.77) |
|  | DT | 0.72 (0.71 - 0.73) | 0.70 (0.69 - 0.72) | 0.72 (0.70 - 0.74) | 0.71 (0.70 - 0.73) |
|  | RF | 0.78 (0.77 - 0.80) | 0.78 (0.77 - 0.80) | 0.75 (0.73 - 0.76) | 0.76 (0.75 - 0.78) |
|  | SVM | 0.78 (0.76 - 0.79) | 0.78 (0.76 - 0.80) | 0.75 (0.73 - 0.76) | 0.76 (0.75 - 0.78) |
|  | XGBoost | 0.77 (0.76 - 0.79) | 0.77 (0.75 - 0.79) | 0.75 (0.73 - 0.77) | 0.76 (0.75 - 0.77) |
| **Demographics** | LR | 0.56 (0.54 - 0.57) | 0.55 (0.52 - 0.57) | 0.39 (0.37 - 0.42) | 0.46 (0.44 - 0.48) |
|  | DT | 0.55 (0.54 - 0.57) | 0.55 (0.52 - 0.57) | 0.39 (0.37 - 0.41) | 0.45 (0.43 - 0.47) |
|  | RF | 0.55 (0.54 - 0.56) | 0.54 (0.51 - 0.56) | 0.41 (0.39 - 0.43) | 0.47 (0.45 - 0.49) |
|  | SVM | 0.53 (0.51 - 0.54) | 0.56 (0.49 - 0.62) | 0.06 (0.05 - 0.07) | 0.11 (0.09 - 0.13) |
|  | XGBoost | 0.56 (0.54 - 0.57) | 0.56 (0.53 - 0.58) | 0.39 (0.37 - 0.41) | 0.46 (0.44 - 0.48) |
| **All features** | LR | 0.75 (0.74 - 0.76) | 0.73 (0.72 - 0.75) | 0.76 (0.74 - 0.78) | 0.75 (0.73 - 0.76) |
|  | DT | 0.71 (0.70 - 0.72) | 0.70 (0.68 - 0.71) | 0.70 (0.68 - 0.72) | 0.70 (0.68 - 0.72) |
|  | RF | 0.77 (0.76 - 0.78) | 0.78 (0.76 - 0.79) | 0.73 (0.71 - 0.75) | 0.75 (0.74 - 0.77) |
|  | SVM | 0.76 (0.75 - 0.77) | 0.76 (0.74 - 0.78) | 0.72 (0.71 - 0.74) | 0.74 (0.73 - 0.75) |
|  | XGBoost | 0.77 (0.76 - 0.78) | 0.77 (0.75 - 0.78) | 0.75 (0.73 - 0.77) | 0.76 (0.74 - 0.77) |

Values are presented as mean (95% CI). AUROC indicates area under the receiver operating characteristic curve; BP, BP; DT, decision tree; LR, logistic regression; RF, Random Forest; SVM, support vector machine.

### Demographic features only (group 2)

Models utilizing demographic factors exclusively showed markedly limited predictive capability (**Table 2**). Accuracy and precision metrics were only marginally superior to random chance (0.5), with accuracy reaching maximum values of 0.56 for XGBoost. Recall and F1-scores were considerably lower, indicating particular difficulty in identifying patients with uncontrolled BP using demographic information alone. XGBoost achieved the highest accuracy (0.56) and precision (0.56), while RF demonstrated the highest recall (0.41) and F1-score (0.47), though these values remained substantially below clinically useful thresholds. The AUROC curves for demographic-only models (**Figure 1B**) confirmed overall poor discriminative performance, with XGBoost showing modest superiority but still limited clinical utility (AUROC approximately 0.60).

**Fig 1B.**
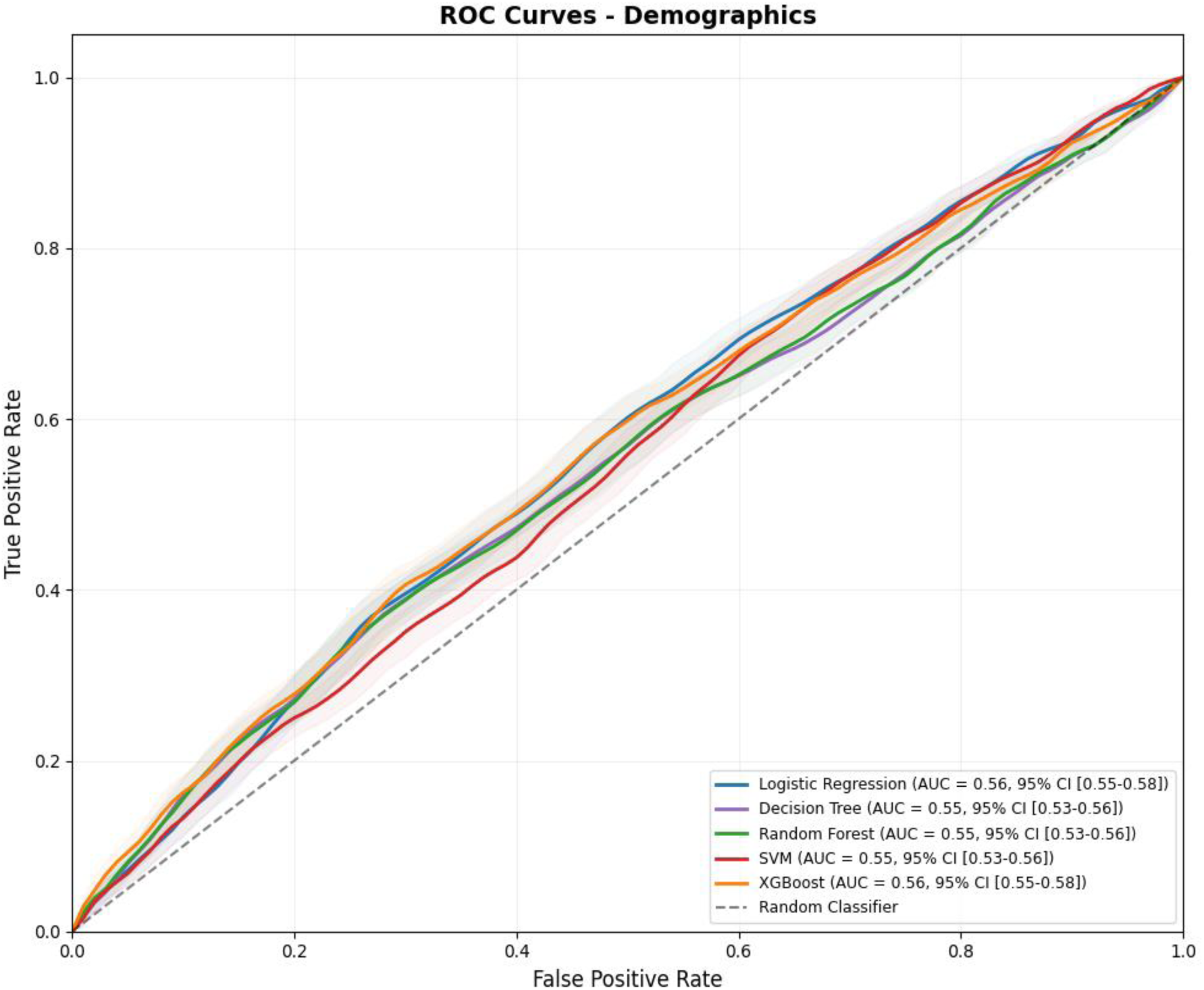
AUROC among five models of demographics (hypertension cohort)

### Combined BP history and demographic features (group 3)

When combining BP history with demographic characteristics, model performance improved modestly compared to BP history alone for some algorithms but did not substantially exceed the performance achieved with BP features exclusively (**Table 2**). RF achieved the highest accuracy (0.77) and precision (0.78), while Logistic Regression demonstrated the highest recall (0.76). The F1-scores were comparable across LR, RF, and XGBoost, ranging from 0.75 to 0.76. The AUROC curves for combined feature models (**Figure 1C**) indicated that RF (AUROC = 0.87) and XGBoost (AUROC = 0.87) demonstrated superior and nearly equivalent discriminative performance. Notably, the combined feature models did not meaningfully improve upon BP history-only models for RF (0.88 vs 0.87), suggesting that demographic features contributed minimal additional predictive information beyond what was captured by longitudinal BP patterns. Based on comprehensive evaluation of performance metrics across all three feature groups and five modeling algorithms, RF emerged as the optimal model, demonstrating consistent superior performance, particularly with BP history features. The finding that BP history alone achieved comparable or superior performance to combined features underscores the primacy of physiological measurements in predicting control status.

**Fig 1C.**
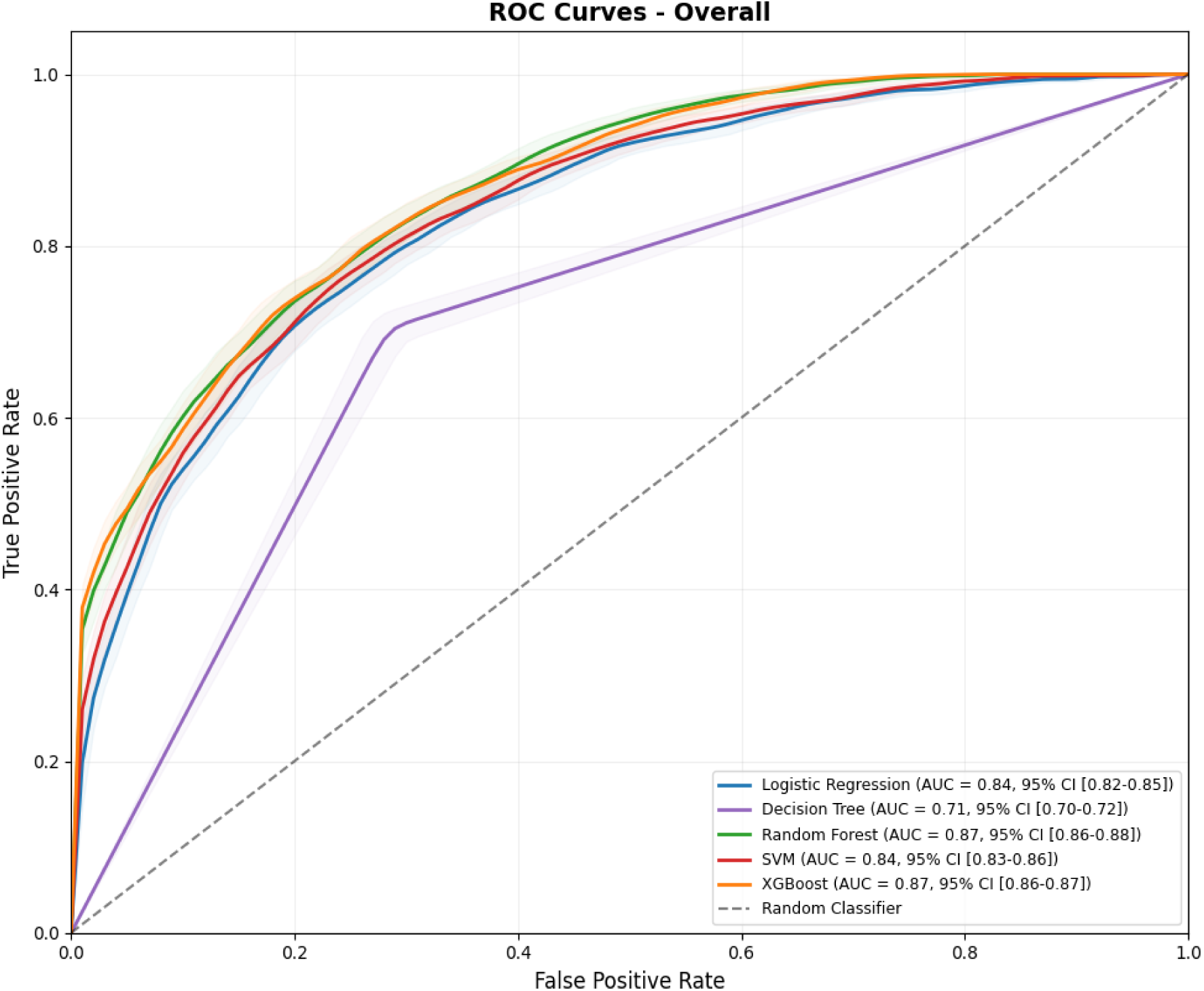
AUROC among five models of all features (hypertension cohort)

### Model interpretation: hypertension cohort

To elucidate which specific factors most substantially influenced predictions in the best-performing model, we conducted SHAP analysis on the RF model incorporating all available features. The feature importance ranking based on mean absolute SHAP values (**Figure 2A**) revealed a clear hierarchy of predictive factors. Mean systolic BP emerged as the single most influential predictor, followed sequentially by maximum systolic BP, maximum diastolic BP, and mean diastolic BP. This ranking demonstrates that both average BP levels and extreme BP values (particularly systolic measurements) contain critical prognostic information. The SHAP summary plot (**Figure 2B**) provides additional insights into the directional relationships between feature values and predicted outcomes. The horizontal axis represents the SHAP value’s contribution to the model output, with negative values (left side) indicating contributions toward predicting controlled BP and positive values (right side) indicating contributions toward predicting uncontrolled BP. The color coding indicates feature magnitude, with red representing high feature values and blue representing low values for each observation.

**Figure 2.**
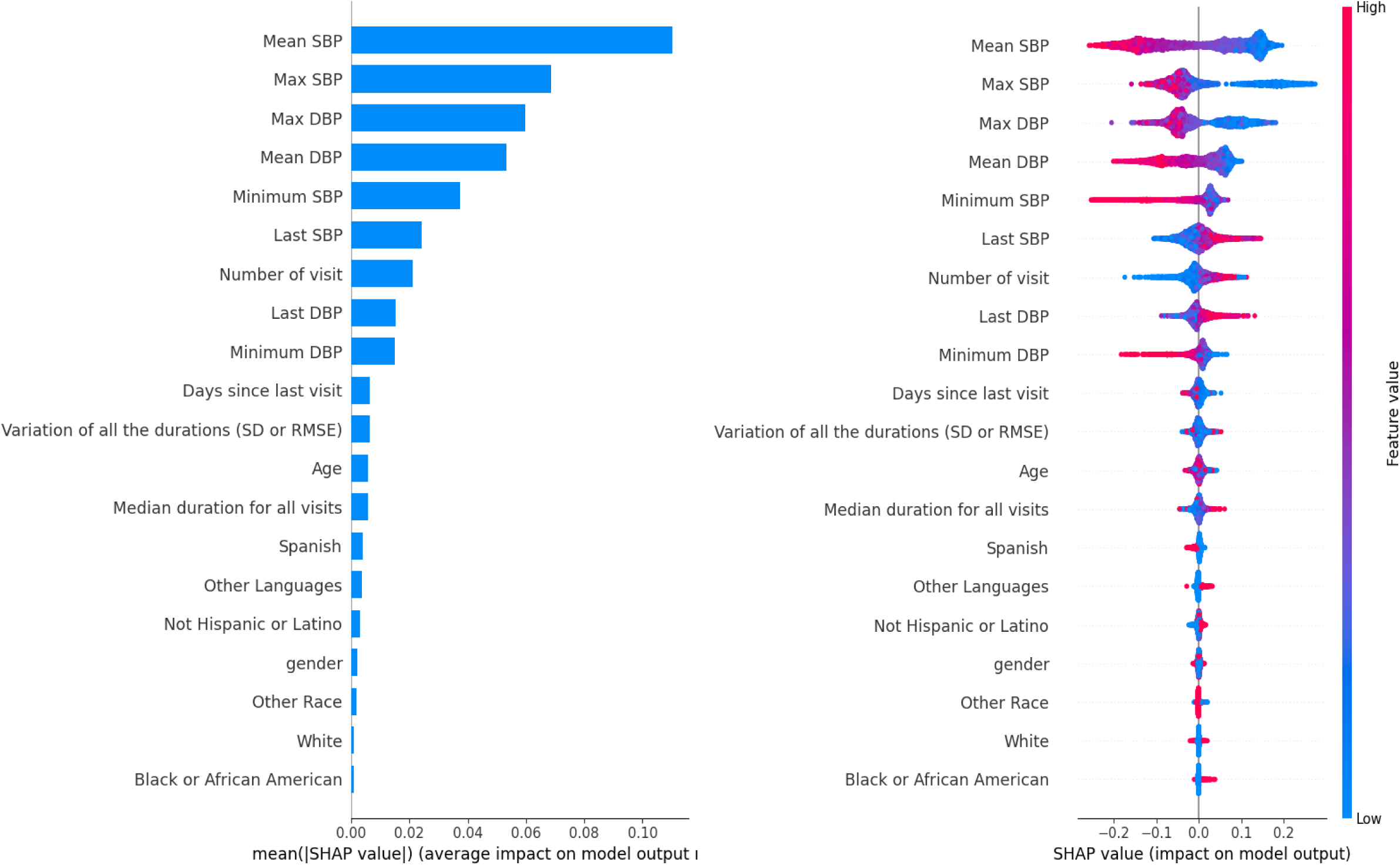
(**A**) Importance ranking of features (hypertension cohort); **(B)** Distribution of impacts of each feature (hypertensive cohort)

The summary plot reveals clear patterns: Lower mean systolic BP values (blue points) are consistently associated with negative SHAP values, indicating strong contributions toward predicting controlled hypertension. Conversely, higher mean systolic BP values (red points) yield positive SHAP values, pushing predictions toward the uncontrolled category. Similarly, elevated maximum systolic BP values (red) consistently generate positive SHAP values, indicating adverse impact on control probability. These relationships align with established clinical knowledge that sustained elevation and extreme BP spikes both indicate inadequate control and elevated cardiovascular risk. Interestingly, while demographic features were included in this comprehensive model, their SHAP values were substantially smaller in magnitude, appearing lower in the importance ranking.

### Hear failure cohort

Among 1,363 patients with documented heart failure diagnoses, 1,137 (83.4%) had completed at least two clinical visits and comprised the final analytical cohort for heart failure analyses. The cohort was nearly evenly divided by sex, with 579 females (50.92%) and 558 males (49.08%). Female patients were significantly older (76.98 ± 15.13 years) compared to male patients (73.25 ± 14.75 years; P<0.001), consistent with known sex differences in heart failure epidemiology and the higher prevalence of heart failure with preserved ejection fraction in older women. Demographic characteristics showed some notable patterns, though not all differences reached statistical significance (**Table 3**). Ethnicity distribution did not differ significantly between sexes (P=0.099), though a numerically higher proportion of males (24.37%) identified as Hispanic or Latino compared to females (18.65%), while 58.03% of females and 55.91% of males identified as not Hispanic or Latino. Among patients reporting specific racial categories, White was the most common (19.69% of females, 17.74% of males), followed by Black or African American (6.22% of females, 4.66% of males) and Asian (0.86% of females, 1.79% of males). Preferred language showed statistically significant differences between sexes (P=0.008). English was the most common language for both groups (46.11% of females, 46.77% of males). However, males demonstrated substantially higher Spanish language preference (18.46%) compared to females (11.92%), while females more frequently spoke languages categorized as “Other” (41.97% vs 34.77% for males).

**Table 3.** Demographic differences by sex (heart failure cohort) Data are presented as No. (%) unless otherwise indicated.

|  | **Total (N=1137)** | **Female (N=579)** | **Male (N=558)** | **p-value** |
| --- | --- | --- | --- | --- |
| **Age, Mean (SD)** | 75.15 (15.05) | 76.98 (15.13) | 73.25 (14.75) | **<0.001** |
| **Ethnicity** |  |  |  | 0.099 |
| Hispanic or Latino | 244 (21.46%) | 108 (18.65%) | 136 (24.37%) |  |
| Not Hispanic or Latino | 648 (56.99%) | 336 (58.03%) | 312 (55.91%) |  |
| Other Ethnicity | 245 (21.55%) | 135 (23.32%) | 110 (19.71%) |  |
| **Race** |  |  |  | 0.413 |
| Asian | 15 (1.32%) | 5 (0.86%) | 10 (1.79%) |  |
| Black or African American | 62 (5.45%) | 36 (6.22%) | 26 (4.66%) |  |
| White | 213 (18.73%) | 114 (19.69%) | 99 (17.74%) |  |
| Other | 847 (74.49%) | 424 (73.23%) | 423 (75.81%) |  |
| **Language** |  |  |  | **0.008** |
| English | 528 (46.44%) | 267 (46.11%) | 261 (46.77%) |  |
| Spanish | 172 (15.13%) | 69 (11.92%) | 103 (18.46%) |  |
| Other | 437 (38.43%) | 243 (41.97%) | 194 (34.77%) |  |

Data are presented as No. (%) unless otherwise indicated.

BP and visit characteristics for the heart failure cohort (**Supplemental Table 2**) revealed patterns somewhat similar to the hypertension cohort but with notable differences reflecting the distinct pathophysiology and clinical management of heart failure. The mean of average systolic BP was 132.96 mmHg, slightly lower than the hypertension cohort, while mean diastolic BP was 77.55 mmHg, also lower. The mean of maximum systolic BP was 156.50 mmHg, with mean minimum diastolic BP of 64.35 mmHg, the latter being notably lower than in the hypertension cohort and potentially reflecting the hemodynamic characteristics of heart failure patients. Patients in the heart failure cohort completed an average of 13.45 visits, higher than the hypertension cohort, likely reflecting more intensive monitoring requirements for heart failure management. The mean duration since last visit was 106.52 days, with median inter-visit interval of 54.00 days.

### Model performance: heart failure cohort BP history features only (group 1)

For the heart failure cohort with BP history features exclusively, RF achieved the highest accuracy at 0.86 and the highest precision at 0.73 (**Table 4**). However, recall and F1-score metrics were substantially lower than in the hypertension cohort and in some cases lower than expected by chance, likely attributable to the considerably smaller sample size and pronounced class imbalance in the heart failure cohort. Decision Tree showed inferior performance compared to other algorithms, consistent with its behavior in the hypertension cohort. The AUROC curves (**Figure 3A**) indicated that RF demonstrated the best discriminative performance among all models for the heart failure cohort using BP history alone, though precise AUROC values should be interpreted cautiously given the limited sample size.

**Fig 3A.**
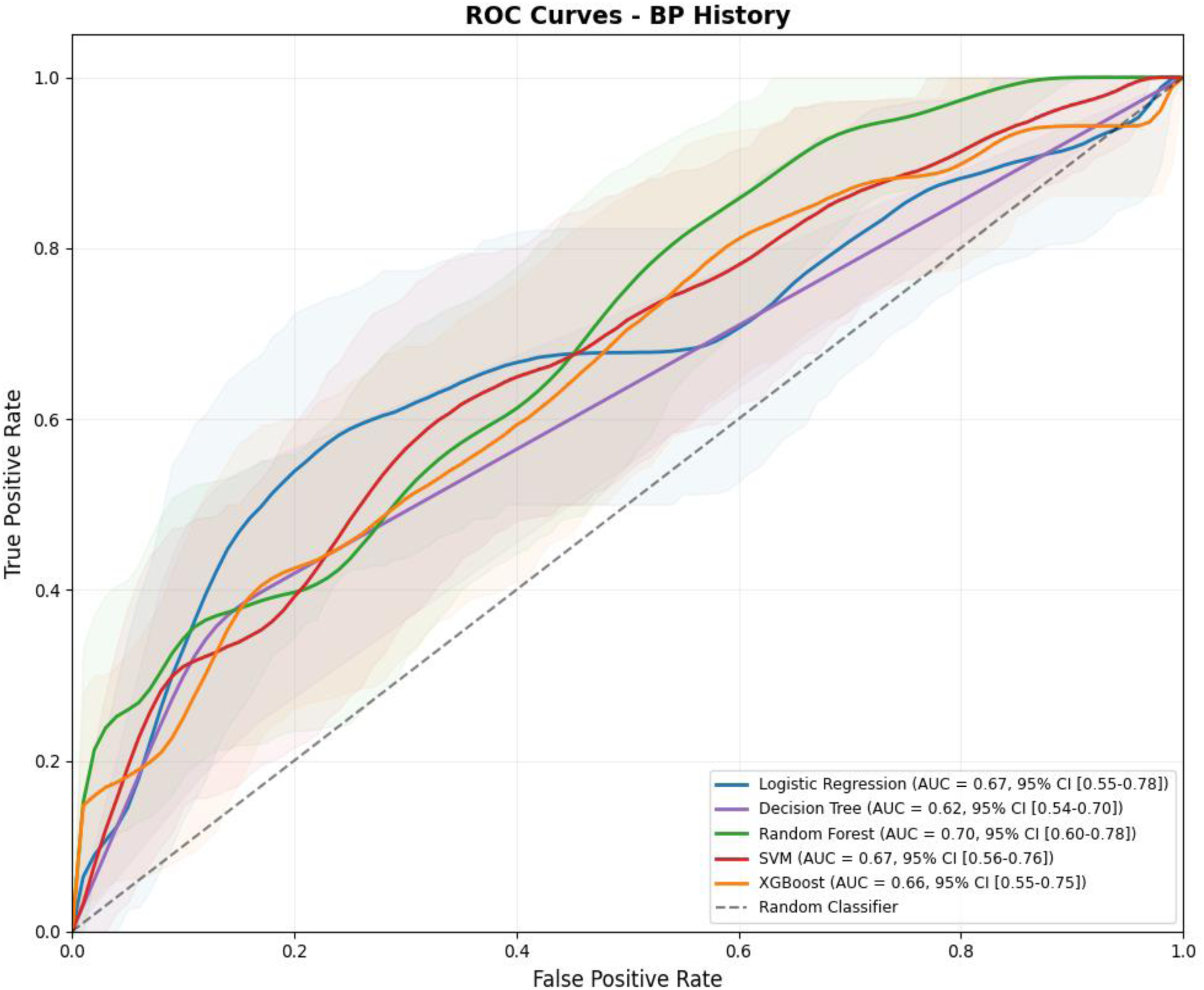
AUROC among five models of BP history (heart failure cohort)

**Table 4.** Models’ performance of three feature groups (heart failure cohort)

| Feature group | Model | Accuracy | Precision | Recall | F1-score |
| --- | --- | --- | --- | --- | --- |
| **BP history** | LR | 0.85 (0.80 - 0.90) | 0.59 (0.00 - 1.00) | 0.03 (0.00 - 1.00) | 0.05 (0.00 - 1.00) |
|  | DT | 0.80 (0.75 - 0.84) | 0.35 (0.21 - 0.51) | 0.37 (0.21 - 0.53) | 0.36 (0.21 - 0.49) |
|  | RF | 0.86 (0.81 - 0.90) | 0.73 (0.00 - 1.00) | 0.09 (0.00 - 1.00) | 0.15 (0.00 - 0.31) |
|  | SVM | 0.85 (0.80 - 0.89) | 0.00 (0.00 - 0.00) | 0.00 (0.00 - 0.00) | 0.00 (0.00 - 0.00) |
|  | XGBoost | 0.84 (0.79 - 0.89) | 0.45 (0.17 - 0.73) | 0.17 (0.04 - 0.30) | 0.24 (0.07 - 0.41) |
| **Demographics** | LR | 0.87 (0.83 - 0.91) | 0.65 (0.42 - 0.86) | 0.37 (0.21 - 0.53) | 0.47 (0.29 - 0.62) |
|  | DT | 0.84 (0.79 - 0.88) | 0.47 (0.30 - 0.64) | 0.45 (0.28 - 0.63) | 0.46 (0.30 - 0.50) |
|  | RF | 0.84 (0.79 - 0.89) | 0.47 (0.29 - 0.63) | 0.43 (0.27 - 0.59) | 0.44 (0.29 - 0.58) |
|  | SVM | 0.85 (0.80 - 0.89) | 0.00 (0.00 - 0.00) | 0.00 (0.00 - 0.00) | 0.00 (0.00 - 0.00) |
|  | XGBoost | 0.85 (0.80 - 0.90) | 0.50 (0.33 - 0.68) | 0.46 (0.30 - 0.63) | 0.48 (0.32 - 0.62) |
| **All features** | LR | 0.86 (0.82 - 0.90) | 0.57 (0.37 - 0.78) | 0.37 (0.22 - 0.54) | 0.45 (0.29 - 0.60) |
|  | DT | 0.84 (0.79 - 0.88) | 0.48 (0.31 - 0.64) | 0.43 (0.27 - 0.59) | 0.45 (0.30 - 0.58) |
|  | RF | 0.90 (0.86 - 0.94) | 0.76 (0.59 - 0.92) | 0.55 (0.38 - 0.71) | 0.63 (0.48 - 0.77) |
|  | SVM | 0.90 (0.86 - 0.93) | 0.80 (0.61 - 0.95) | 0.46 (0.29 - 0.63) | 0.58 (0.41 - 0.72) |
|  | XGBoost | 0.87 (0.84 - 0.93) | 0.66 (0.48 - 0.84) | 0.54 (0.38 - 0.71) | 0.59 (0.44 - 0.73) |

Values are presented as mean (95% CI). Wide confidence intervals for some metrics reflect small sample size and class imbalance. AUROC indicates area under the receiver operating characteristic curve; BP, BP; DT, decision tree; LR, logistic regression; RF, Random Forest; SVM, support vector machine.

### Demographic features only (group 2)

Models using demographic features alone for the heart failure cohort exhibited a paradoxical pattern: high accuracy but poor precision, recall, and F1-scores, all falling below 0.5 (**Table 4**). This pattern suggests that models were primarily predicting the majority class (controlled BP) for most observations, achieving high overall accuracy due to class imbalance but failing to adequately identify patients with uncontrolled BP. Logistic Regression achieved the best balance with precision of 0.65 and recall of 0.37, though F1-score remained modest at 0.47. The AUROC curves (**Figure 3B**) revealed that Logistic Regression demonstrated the best discriminative performance among demographic-only models, followed by RF and XGBoost.

**Fig 3B.**
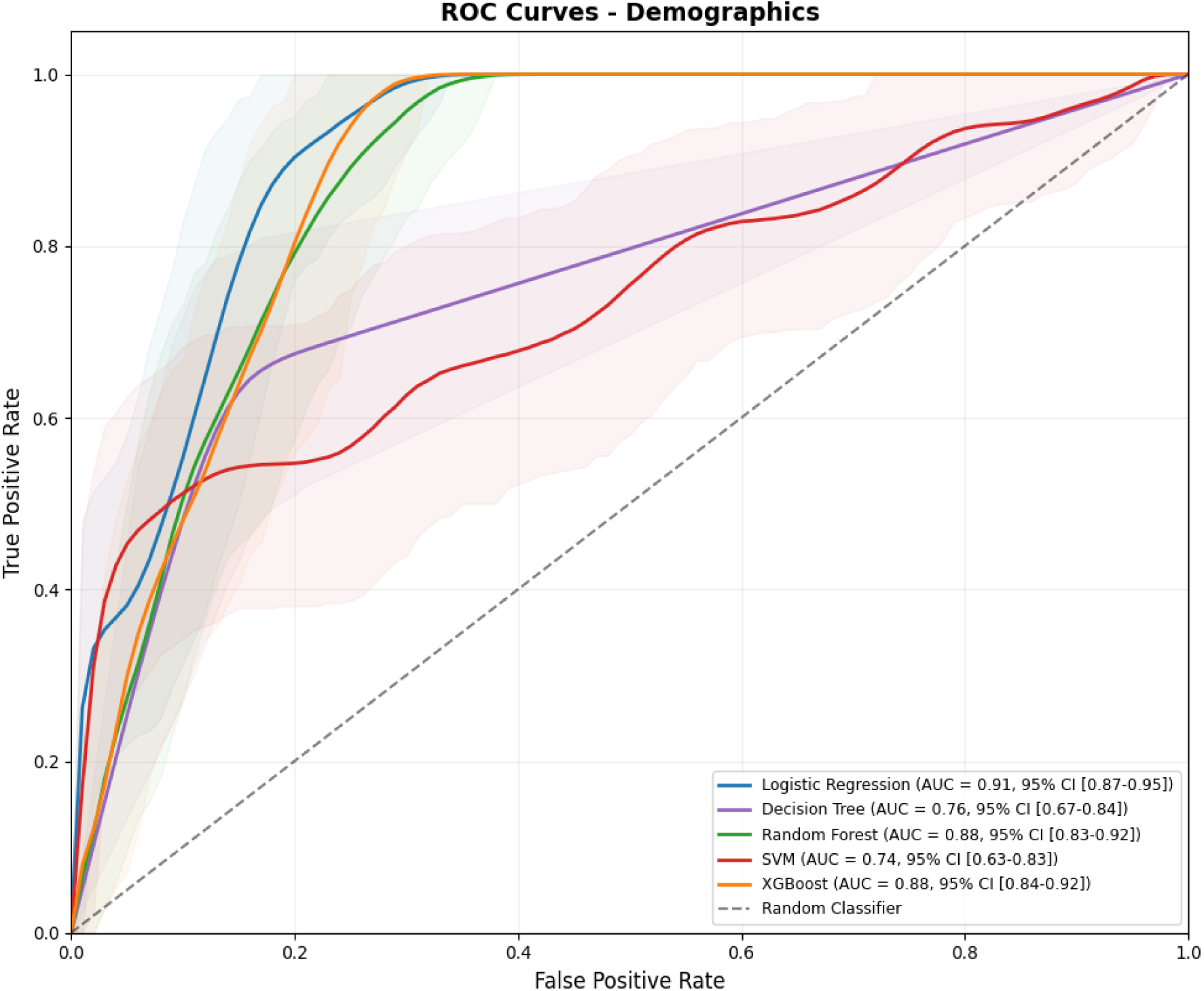
AUROC among five models of demographics (heart failure cohort)

### Combined BP history and demographic features (group 3)

With combined features, model performance improved substantially for most algorithms (**Table 4**). RF and SVM achieved the highest accuracy at 0.90, representing notable improvement over feature groups used in isolation. RF demonstrated precision of 0.76, recall of 0.55, and F1-score of 0.63, indicating more balanced performance across metrics compared to individual feature groups. The AUROC curves (**Figure 3C**) revealed that RF achieved excellent discrimination with an AUROC of 0.93, representing the highest performance observed across all models and cohorts in this study. XGBoost also demonstrated strong performance with an AUROC of 0.92. These results indicate that for heart failure patients, the combination of BP history and demographic information provides complementary predictive value, unlike the hypertension cohort where demographic features contributed minimally beyond BP history alone. Based on comprehensive performance evaluation across feature groups in the heart failure cohort, RF emerged as the optimal modeling approach, with combined BP history and demographic features providing the best predictive framework.

**Fig 3C.**
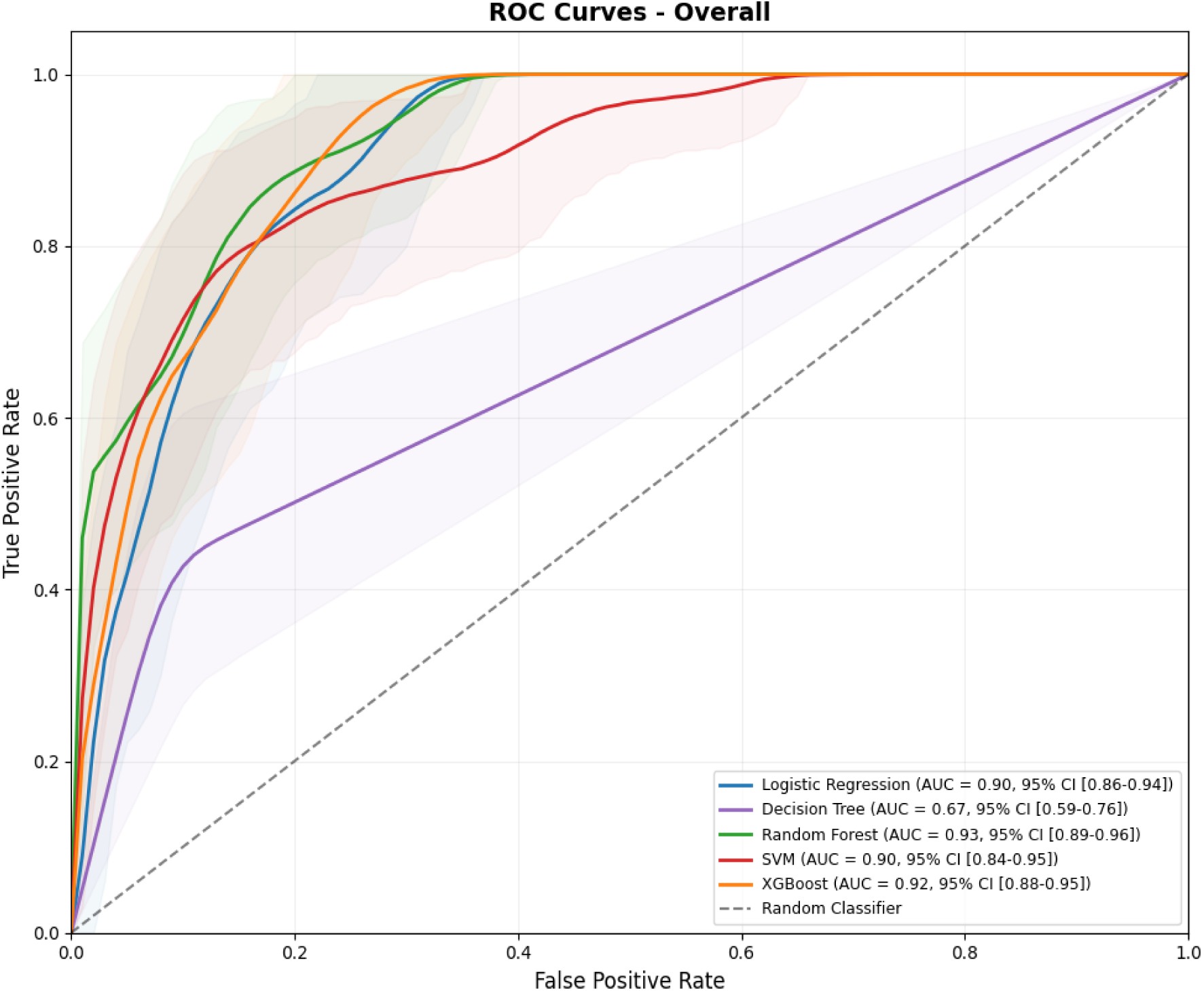
AUROC among five models of all features (heart failure cohort)

### Model interpretation: heart failure cohort

SHAP analysis of the RF model incorporating all features for the heart failure cohort revealed a markedly different pattern of influential predictors compared to the hypertension cohort (**Figure 4A**). The feature importance ranking identified language preference (specifically “Other Languages”) as the most influential predictor, followed by race categories (”Other Race,” “White,” and “Black”), median duration for all visits, and days since last visit. This ranking pattern diverges substantially from the hypertension cohort, where BP measurements dominated the importance hierarchy. The SHAP summary plot (**Figure 4B**) provides insights into directional relationships between features and predicted outcomes in the heart failure cohort. Patients speaking uncommon languages (categorized as “Other Languages”) showed a tendency toward higher heart failure risk in model predictions, potentially reflecting healthcare access barriers, health literacy challenges, or cultural factors influencing disease management and outcomes. Patients with longer median visit durations demonstrated higher predicted heart failure risk, which may indicate more complex clinical presentations requiring extended consultations, greater disease severity, or presence of multiple comorbidities necessitating comprehensive care.

**Figure 4.**
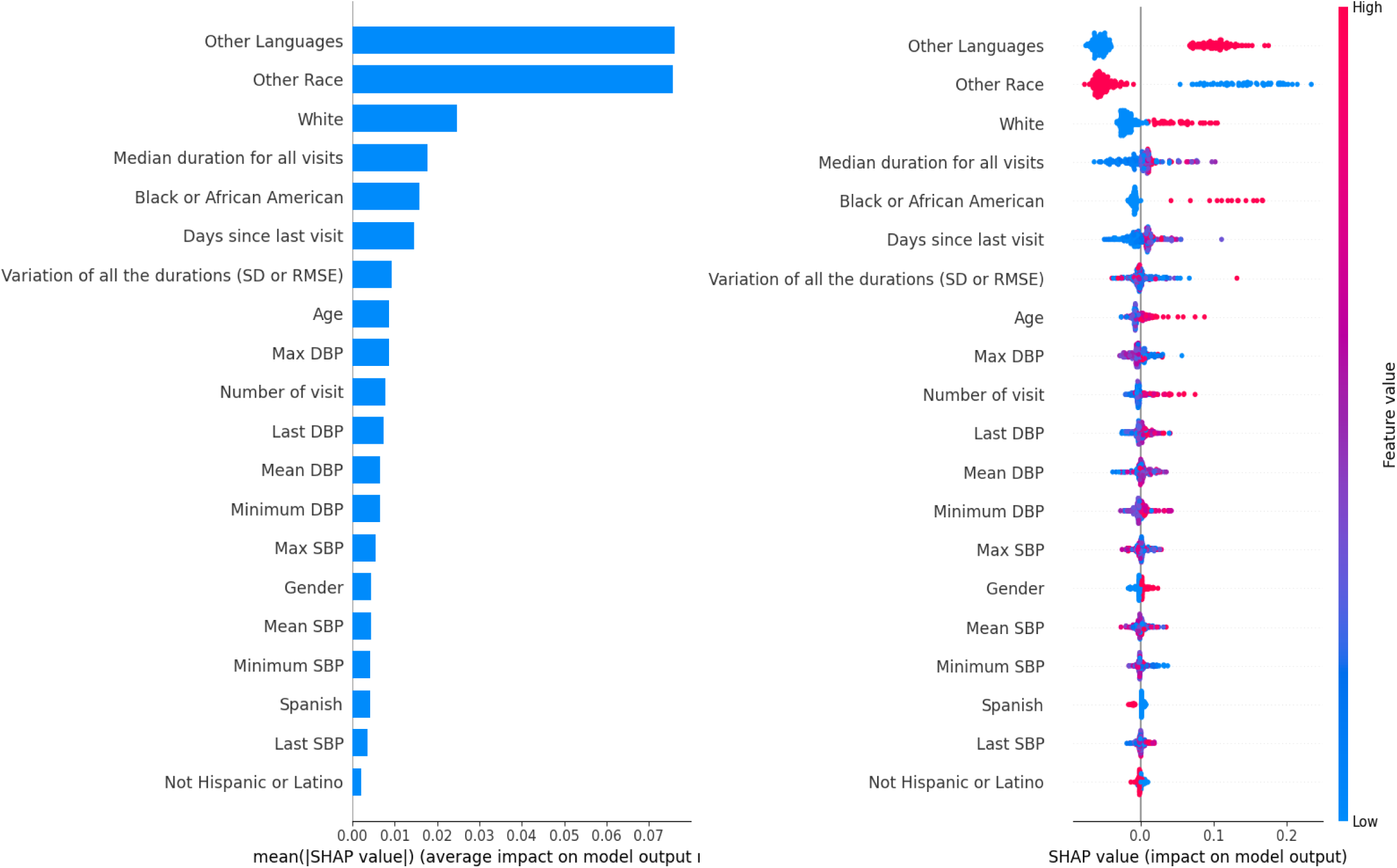
(**A**) Importance ranking of features (heart failure cohort); **(B)** Distribution of impacts of each feature ((heart failure cohort)

The prominent role of demographic and healthcare utilization factors in the heart failure cohort, contrasting with the primacy of BP measurements in the hypertension cohort, likely reflects fundamental differences in disease pathophysiology, outcome definitions, and cohort characteristics. Heart failure is a more heterogeneous syndrome with multiple etiologies and phenotypes, where social determinants of health and healthcare access patterns may play more substantial roles in outcomes.

## DISCUSSION

### Principal Findings

This study demonstrates that machine learning models, particularly RF algorithms, can effectively predict hypertension control status using routinely collected clinical data. Our most significant finding is that longitudinal BP history substantially outperformed demographic characteristics in predicting BP control, achieving an AUROC of 0.88 with BP features alone compared to only 0.60 with demographic features exclusively. The combination of BP history and demographics yielded an AUROC of 0.87, only marginally different from BP history alone, indicating that demographic factors contribute minimal incremental predictive value beyond physiological measurements. SHAP interpretability analysis revealed that mean systolic BP, maximum systolic BP, and BP extremes (maximum diastolic BP) were the most influential predictors of control status. This finding aligns with and extends previous epidemiological research demonstrating that both sustained BP elevation and visit-to-visit variability contribute independently to cardiovascular risk. For the heart failure cohort, RF achieved an AUROC of 0.93 with combined features, though the feature importance pattern differed markedly, with demographic and healthcare utilization factors playing more prominent roles.

### Model selection rationale and performance comparison

RF consistently demonstrated superior or equivalent performance compared to alternative machine learning algorithms across both cohorts and all feature configurations. This consistent advantage likely stems from several algorithmic characteristics that are particularly well-suited to this prediction task. RF’s ensemble approach, which aggregates predictions from hundreds of independently trained decision trees using bootstrap sampling and random feature selection, provides robust protection against overfitting while effectively capturing complex, non-linear relationships between predictors and outcomes.

Unlike gradient boosting methods such as XGBoost, which sequentially construct trees where each attempts to correct previous errors and may be more susceptible to overfitting in smaller datasets or in the presence of noisy labels, RF’s bagging (bootstrap aggregating) methodology provides more conservative predictions by averaging across diverse tree structures. This characteristic proved particularly valuable in our study given the moderate sample size of the hypertension cohort and especially the limited sample size of the heart failure cohort. Additionally, RF naturally handles mixed data types (continuous BP measurements, categorical demographic variables) and missing values without requiring extensive preprocessing, enhancing its practical utility for real-world electronic health record data.

The superior performance of BP history over demographic characteristics carries important implications for clinical risk stratification and resource allocation. This finding suggests that hypertension control is primarily determined by modifiable physiological factors and treatment effectiveness rather than fixed demographic characteristics. From a clinical and health equity perspective, this is encouraging because it implies that quality care and adequate treatment can overcome demographic disparities in achieving BP control.[14] However, it simultaneously highlights that demographic factors likely influence hypertension development and healthcare access rather than control once diagnosis and treatment are established, emphasizing the need for upstream interventions addressing social determinants of health.[15]

### Comparison with previous literature

Our findings complement and extend previous research examining relationships between healthcare visit patterns and hypertension outcomes. Guthmann et al. demonstrated that shorter visit intervals were associated with improved BP control, with statistically significant correlations between visit frequency and changes in both systolic and diastolic BP.[16] Montayre et al. similarly found that increased utilization of nurse visits was associated with better BP control, particularly among females with treated hypertension.[17] These studies established the general principle that more frequent healthcare contact improves outcomes, likely through enhanced medication titration, improved adherence, and earlier identification of control problems.[18]

In contrast, Kim et al. reported that longer visit intervals (3-6 months) did not significantly increase risk of major adverse cardiovascular events or all-cause mortality in hypertensive patients, suggesting that extended follow-up intervals after medication stabilization may represent a cost-effective approach for selected patients.[19] This apparent contradiction with studies showing benefits of frequent visits likely reflects differences in patient populations (stable vs. newly diagnosed or uncontrolled patients) and outcome definitions (BP control vs. major adverse events). Our study extends this literature by systematically quantifying how comprehensive visit history characteristics, including visit frequency, inter-visit intervals, and interval variability, contribute to predicting BP control status. While visit-related features appeared in our models, they were substantially less influential than BP measurements themselves according to SHAP analysis. This suggests that what happens during visits (achieving BP control) matters more than visit frequency per se, though adequate visit frequency is presumably necessary for monitoring and treatment optimization.[20]

Higashiyama et al. focused on self-reported hypertension and its relationship with cardiovascular disease (CVD) mortality, using the Cox proportional hazard model to show that self-reported hypertension significantly increased CVD mortality risk, even in the absence of confirmative hypertension [21]. Rothwell et al. conducted landmark research identifying visit-to-visit systolic BP variability and maximum systolic BP as strong predictors of stroke risk, independent of mean BP, with particularly pronounced effects in younger patients and those with lower mean systolic BP.[22] Our SHAP analysis strongly corroborates these findings in a different outcome context (BP control status rather than stroke risk), demonstrating that maximum BP values substantially contribute to predictions even after accounting for mean BP. This convergence between traditional epidemiological approaches and modern machine learning methods strengthens the evidence base for incorporating BP variability metrics into routine clinical assessment and risk stratification.[23]

Our study’s findings can be contextualized within the broader landscape of machine learning applications for hypertension-related predictions. Kawasoe et al. developed a risk prediction score for hypertension incidence using logistic regression, achieving an AUROC of 0.76, while Deng et al. created a nomogram for hypertension prediction with an AUROC of 0.772.[24, 25] These studies focused on predicting hypertension development rather than control status and relied on traditional statistical approaches, which while interpretable, may not fully capture non-linear relationships in data. Feng et al. applied more advanced machine learning techniques including random survival forests and neural multi-task logistic regression to predict cardiovascular hospitalizations in hypertensive patients, achieving concordance indices up to 0.8149 [26]. Wang et al. employed 5-fold cross-validation frameworks and demonstrated the utility of frequent health checkups in improving hypertension predictions.[27] Both studies emphasized the importance of comprehensive datasets with detailed longitudinal features, an approach mirroring our incorporation of extensive BP history statistics.

Several studies have applied machine learning specifically to predict hypertension control outcomes with performance metrics.[28, 29] Xiang et al. predicted disability risk in elderly adults with hypertension using multiple algorithms including logistic regression, gradient boosting, and deep neural networks, achieving AUROCs up to 0.759.[30] Mroz et al. used XGBoost to predict hypertension control, achieving an AUC of 0.76 [31]. Nguyen et al. developed models for sustained uncontrolled hypertension and hypertensive crisis within one year. Our RF model’s AUROC of 0.88 represents superior performance compared to these studies, potentially attributable to our comprehensive feature engineering incorporating detailed BP variability metrics and visit pattern characteristics.

Recent advances in deep learning have demonstrated potential for leveraging temporal and longitudinal data for cardiovascular predictions. Yun et al. utilized temporal fusion transformers to predict intradialytic hypotension and hypertension, achieving impressive AUROCs exceeding 0.95.[32] Datta et al. demonstrated LSTM network superiority for predicting hypertension onset with an AUC of 0.94.[33] While these deep learning approaches achieve exceptional performance, they typically sacrifice interpretability, which remains crucial for clinical adoption and trust. Our use of SHAP with RF provides an advantageous balance between predictive accuracy and transparency, offering clinically actionable insights about which factors drive predictions for individual patients. Du et al. developed a web-based hypertension risk prediction system using LightGBM, highlighting the importance of user-friendly visualization in translating machine learning outputs into actionable recommendations for patients and clinicians.[34] Similarly, our study’s emphasis on interpretable models supported by SHAP analysis aims to facilitate clinical understanding and potential integration into decision-support systems, recognizing that technical performance alone is insufficient for successful clinical implementation without accompanying interpretability and usability.

In exploring predictors of hypertension and cardiovascular outcomes, traditional statistical analyses have provided valuable insights but remain limited in capturing complex, multidimensional relationships. Kario et al. demonstrated that arterial stiffness (CAVI) and small artery retinopathy independently predict hypertension development, emphasizing the role of structural vascular changes in disease progression [35]. Similarly, Gallagher et al. highlighted the predictive utility of B-type natriuretic peptide (BNP) for cardiovascular events and mortality, showing its superiority over ventricular dysfunction and risk scores like SCORE in stratifying patient risk [36]. Chowdhury et al. identified short-term BP variability, particularly daytime and weighted day–night systolic BP, as significant predictors of mortality in elderly hypertensive patients, underscoring the importance of temporal BP dynamics [37]. These studies effectively addressed critical aspects of hypertension and cardiovascular risk, relying predominantly on specific biomarkers or physiological indices. They inspired the consideration of integrating a broader range of factors, including specific biomarkers, into predictive models.

### Clinical implications and interpretation of key predictors

Our SHAP analysis revealed that mean systolic BP emerged as the single most influential predictor of future control status, followed closely by maximum systolic BP and maximum diastolic BP. These findings carry several important clinical implications for hypertension management strategies. The prominence of mean systolic BP reflects its role as an integrative measure of overall BP burden over time. Sustained elevation in average systolic BP indicates inadequate therapeutic response or poor medication adherence, both of which require clinical attention. This finding supports current guideline emphasis on achieving and maintaining systolic BP targets, as even modest sustained elevations substantially increase cardiovascular risk.

Equally important is the strong influence of maximum BP values, which capture BP variability and extreme measurements. The clinical significance of BP variability has been increasingly recognized in cardiovascular medicine, with visit-to-visit variability identified as an independent predictor of stroke, myocardial infarction, and cardiovascular mortality. Maximum systolic BP likely represents both true physiological variability (potentially driven by arterial stiffness, autonomic dysfunction, or environmental stressors) and measurement variability (related to technique, equipment, or timing of measurements). Regardless of mechanism, our findings suggest that even transient BP spikes warrant clinical attention and may indicate inadequate control even when average BP appears acceptable.

From a practical clinical perspective, these findings emphasize the importance of comprehensive BP characterization rather than relying on isolated measurements. Current clinical practice often focuses primarily on the most recent BP reading when making treatment decisions. Our results suggest that incorporating historical patterns, particularly tracking mean values over time and identifying concerning maximum values, could enhance clinical decision-making. Electronic health records could be programmed to automatically calculate and display these summary statistics, alerting clinicians to patients with high BP variability or concerning maximum values who might benefit from treatment intensification or more frequent monitoring.

The relatively minor influence of demographic factors (age, race, sex) in our final models, while perhaps surprising given known epidemiological associations between these variables and hypertension prevalence, likely reflects that these factors primarily influence hypertension risk and development rather than control once diagnosis and treatment are established. In other words, while demographic factors help predict who will develop hypertension, physiological measurements better predict whose hypertension will be controlled with treatment. This distinction is crucial for developing targeted interventions: primary prevention efforts should address demographic disparities in hypertension development, while secondary prevention and disease management should focus on optimizing BP control through intensified monitoring and treatment adjustment regardless of demographic characteristics.

### Limitations

This study has several limitations. First, external validation with independent datasets is necessary to assess generalizability across different healthcare settings, patient populations, and geographic regions. Variations in clinical practice patterns, patient demographics, and data quality may affect model transferability. Second, while we compared five common machine learning algorithms, additional methods including deep neural networks, gradient boosting variants, and ensemble meta-learners might yield different performance characteristics. The relatively limited feature set, focused on BP history and basic demographics, may not capture the full complexity of hypertension control. Third, the heart failure cohort’s limited sample size resulted in unstable performance metrics, particularly precision, recall, and F1-scores, due to class imbalance and insufficient positive cases. This limitation is reflected in the wide confidence intervals and occasional extreme values for these metrics. Larger cohorts are needed to develop robust models for heart failure outcomes. Finally, the retrospective observational design cannot establish causality between identified predictors and outcomes. While BP history strongly predicts control status, intervention studies are needed to determine whether targeted modification of these factors improves outcomes.

### Future research

Future investigations should pursue several complementary directions to address current limitations and advance this field toward clinical implementation. First and foremost, rigorous external validation across diverse healthcare settings is essential. Validation studies should encompass academic medical centers, community hospitals, federally qualified health centers, rural clinics, and international healthcare systems to assess model transportability and identify factors limiting generalizability. Cross-institutional validation would reveal whether models trained in one setting maintain performance when applied to patient populations with different demographic compositions, treatment practices, or healthcare access patterns.[38] Geographic validation across regions with varying hypertension prevalence and control rates would further assess robustness. Such validation efforts should systematically examine model performance across patient subgroups defined by age, sex, race, ethnicity, and comorbidity burden to identify potential disparities in prediction accuracy that could perpetuate or exacerbate health inequities if models were deployed clinically.[39]

Second, exploring more sophisticated modeling approaches capable of better capturing temporal dynamics represents a promising avenue. Recurrent neural networks (RNNs), particularly Long Short-Term Memory (LSTM) networks, are specifically designed to process sequential data and might better leverage the temporal ordering of BP measurements. Attention mechanisms could identify which historical time points are most informative for predicting future control. Temporal convolutional networks offer computational efficiency advantages while maintaining ability to process variable-length sequences. However, pursuing these advanced methods must be balanced against interpretability concerns, as more complex models may be harder for clinicians to understand and trust.[40] Developing explainability techniques specifically designed for temporal models, such as temporal attention visualizations or time-series SHAP extensions, would be valuable.

Third, substantially expanding the feature space to incorporate comprehensive clinical and behavioral data would provide more holistic risk assessment.[41] Detailed medication data including specific antihypertensive agents, doses, regimen complexity, and changes over time would illuminate treatment intensification patterns and medication effectiveness.[42] Pharmacy refill data or electronic medication monitoring could provide objective medication adherence metrics.[43] Lifestyle factors measurable through patient-reported outcomes or wearable devices, including physical activity levels, dietary patterns, sodium intake, sleep quality, and stress levels, represent modifiable risk factors that could inform personalized intervention recommendations.[44, 45] Socioeconomic variables including insurance status, income level, educational attainment, and neighborhood deprivation indices could identify patients facing structural barriers to BP control who might benefit from tailored support interventions.[46] Laboratory data including renal function markers (creatinine, estimated glomerular filtration rate), metabolic parameters (glucose, lipids), and inflammatory markers could provide insights into pathophysiological mechanisms and comorbidity burden.

Fourth, integrating real-time data streams from emerging technologies could enable dynamic, continuously updated prediction models. Home BP monitoring devices, increasingly adopted for hypertension management, generate rich time-series data capturing BP patterns under real-world conditions without white-coat effect. Wearable devices capable of continuous or frequent BP measurement (emerging photoplethysmography-based devices, implantable monitors) could provide unprecedented temporal resolution.[47] Electronic medication adherence devices and smartphone applications offer objective adherence data.[48] Real-time integration of such data streams into predictive models could enable truly personalized medicine, with risk assessments continuously updated as new information becomes available and clinical decision support adapting to changing patient status.[49]

Fifth, developing unified, multi-outcome models that simultaneously predict multiple cardiovascular endpoints would better reflect the interrelated nature of cardiovascular diseases. Hypertension, heart failure, coronary artery disease, stroke, and chronic kidney disease share common risk factors and pathophysiological mechanisms, and patients frequently have multiple conditions. Multi-task learning approaches, where a single model is trained to predict multiple related outcomes, could leverage shared risk factors while capturing condition-specific predictors.[50] Such models would provide comprehensive cardiovascular risk assessment from a single integrated evaluation, potentially more efficient and clinically useful than separate disease-specific models. Transfer learning approaches could enable models trained on large hypertension datasets to be adapted for related but less common conditions with limited available data.[51]

Sixth, rigorous prospective implementation studies are essential to demonstrate clinical utility and cost-effectiveness. Cluster randomized controlled trials comparing clinical outcomes in practices using prediction model-guided care versus usual care would provide the highest-level evidence for clinical benefit. Such trials should assess not only BP control rates but also major adverse cardiovascular events, healthcare utilization, costs, and patient-reported outcomes including quality of life and treatment satisfaction. Implementation science approaches are needed to understand barriers and facilitators to clinical adoption, optimize model integration into existing clinical workflows, and develop strategies to minimize alert fatigue while ensuring important predictions trigger appropriate clinical action.[52] Human-centered design methodologies involving clinicians and patients throughout model development and interface design will be crucial for creating decision support tools that are actually used in practice and improve rather than hinder clinical care.

Finally, careful attention to health equity implications throughout model development, validation, and implementation is imperative. Machine learning models can perpetuate or even amplify existing healthcare disparities if trained on biased data or deployed without consideration of differential performance across population subgroups. Future research should explicitly evaluate model performance stratified by race, ethnicity, socioeconomic status, and other social determinants of health. Models should be regularly audited for fairness using established frameworks. Development of techniques to mitigate algorithmic bias while maintaining predictive performance represents an important methodological challenge. Additionally, deployment strategies should consider how to ensure equitable access to model-guided care, avoiding scenarios where sophisticated predictive tools are available only in well-resourced settings, potentially widening existing disparities.

## CONCLUSION

This study demonstrates that machine learning models, particularly RF, can effectively predict hypertension control status using routinely collected clinical data. Longitudinal BP history, especially mean and maximum BP values, substantially outperformed demographic characteristics in predicting control status. The interpretability provided by SHAP analysis offers clinically actionable insights, suggesting that BP variability metrics and historical patterns should be systematically incorporated into hypertension management strategies. These predictive models could serve as valuable decision-support tools, enabling more personalized and proactive care for patients with hypertension and related cardiovascular conditions.

## CONTRIBUTION STATEMENT

JY designed the study, contributed to the data analyses, and the writing of the manuscript. HC contributed to data analyses and the writing of the manuscript. All authors read and approved the final version of the manuscript.

## DATA AVAILABILITY

Datasets are available from the corresponding author on reasonable request and completion of appropriate data sharing agreements.

## Data Availability

All data produced in the present study are available upon reasonable request to the authors

---

**Supplemental Table 1.** BP and visit characteristics: hypertension cohort (n=23,002)

| Variables | Value |
| --- | --- |
| Mean SBP | 135.76 |
| Mean DBP | 81.86 |
| Last SBP | 134.48 |
| Last DBP | 80.72 |
| Minimum SBP | 118.27 |
| Minimum DBP | 70.29 |
| Maximum SBP | 155.69 |
| Maximum DBP | 93.74 |
| Number of visits | 10.61 |
| Days since last visit | 130.11 |
| Median duration for all visits | 65.50 |
| Variation of all the durations (SD or RMSE) | 71.25 |
| Variation of all the durations (SD or RMSE) | 113.48 |

**Supplemental Table 2.**
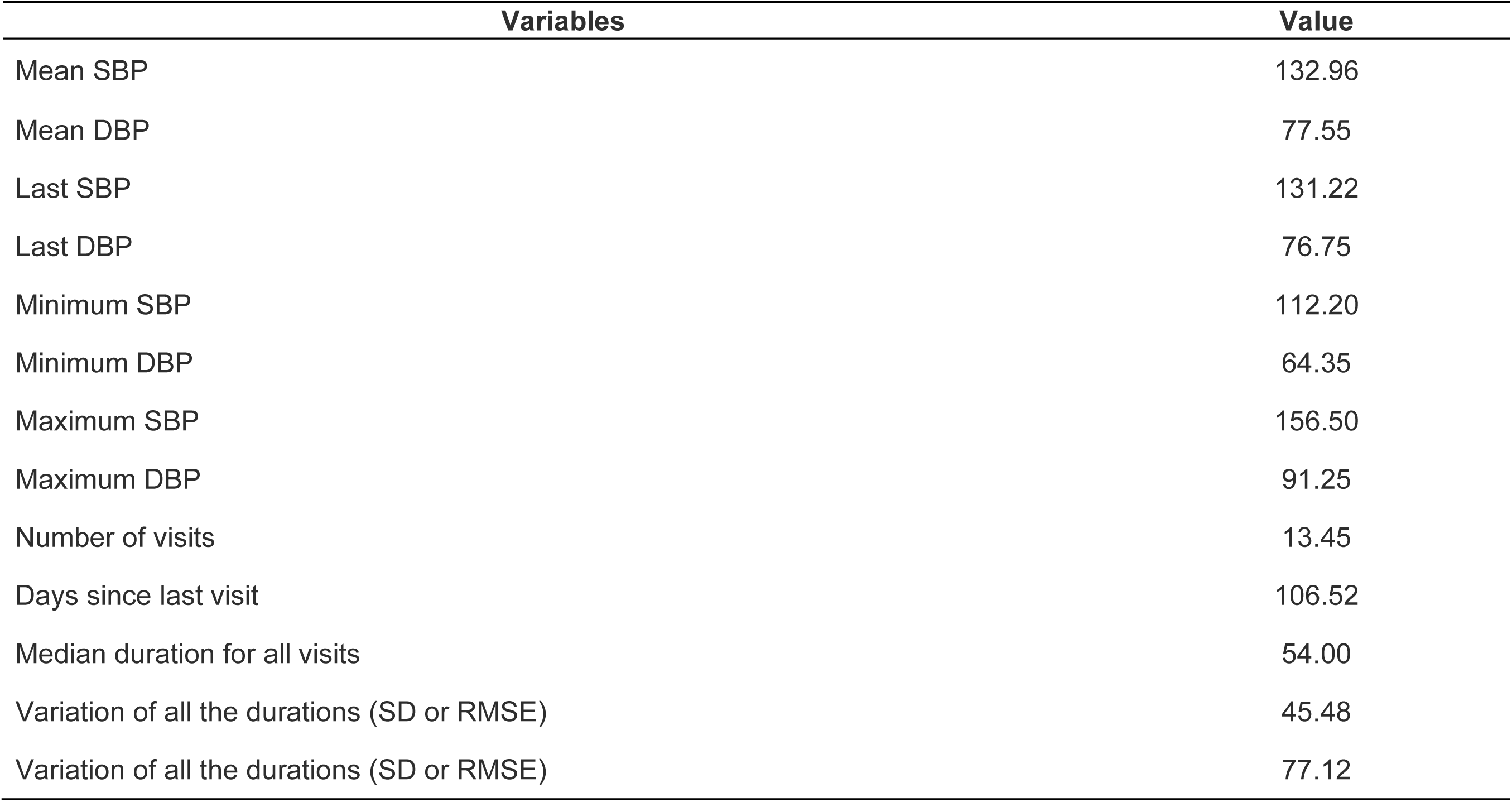
BP and visit characteristics: heart failure cohort (n=1,137)

| Variables | Value |
| --- | --- |
| Mean SBP | 132.96 |
| Mean DBP | 77.55 |
| Last SBP | 131.22 |
| Last DBP | 76.75 |
| Minimum SBP | 112.20 |
| Minimum DBP | 64.35 |
| Maximum SBP | 156.50 |
| Maximum DBP | 91.25 |
| Number of visits | 13.45 |
| Days since last visit | 106.52 |
| Median duration for all visits | 54.00 |
| Variation of all the durations (SD or RMSE) | 45.48 |
| Variation of all the durations (SD or RMSE) | 77.12 |

## REFERENCE

1. Organization, W.H., Hypertension. 2025.

2. Chen, H., et al., Telehealth Utilization and Patient Experiences: The Role of Social Determinants of Health Among Individuals with Hypertension and Diabetes. medRxiv, 2024: p. 2024.08. 01.24311392.

3. Ye, J., et al., Optimizing Longitudinal Retention in Care Among Patients With Hypertension in Primary Healthcare Settings: Findings From the Hypertension Treatment in Nigeria Program. Circulation, 2022. 146(Suppl_1): p. A13217-A13217.

4. Chhabra., P.S.A.M.L., Heart Failure (Congestive Heart Failure). 2025, StatPearls.

5. PREVENTION, U.S.C.F.D.C.A., About Heart Failure. 2024.

6. White-Williams, C., et al., Addressing social determinants of health in the care of patients with heart failure: a scientific statement from the American Heart Association. Circulation, 2020. 141(22): p. e841–e863.

7. Zhang, X., et al., Predicting next-day discharge via electronic health record access logs. Journal of the American Medical Informatics Association, 2021. 28(12): p. 2670–2680.

8. Yan, C., et al., Predicting brain function status changes in critically ill patients via Machine learning. Journal of the American Medical Informatics Association, 2021. 28(11): p. 2412–2422.

9. Li, J., et al., Predicting Mortality in Intensive Care Unit Patients With Heart Failure Using an Interpretable Machine Learning Model: Retrospective Cohort Study. J Med Internet Res, 2022. 24(8): p. e38082.

10. Liu, J., et al., Predicting mortality of patients with acute kidney injury in the ICU using XGBoost model. PLoS One, 2021. 16(2): p. e0246306.

11. Wang, K., et al., Development and validation of a machine learning-based prognostic risk stratification model for acute ischemic stroke. Scientific Reports, 2023. 13(1): p. 13782.

12. Ma, Q., et al., Effect and prediction of physical exercise and diet on blood pressure control in patients with hypertension. Medicine (Baltimore), 2023. 102(50): p. e36612.

13. Kohjitani, H., et al., Recent developments in machine learning modeling methods for hypertension treatment. Hypertension Research, 2024. 47(3): p. 700–707.

14. Ye, J., et al., Identifying Practice Facilitation Delays and Barriers in Primary Care Quality Improvement. The Journal of the American Board of Family Medicine, 2020. 33(5): p. 655–664.

15. Ye, J., Z. Wang, and J. Hai, Social Networking Service, Patient-Generated Health Data, and Population Health Informatics: National Cross-sectional Study of Patterns and Implications of Leveraging Digital Technologies to Support Mental Health and Well-being. Journal of medical Internet research, 2022. 24(4): p. e30898.

16. Guthmann, R., et al., Visit frequency and hypertension. J Clin Hypertens (Greenwich), 2005. 7(6): p. 327–32.

17. Montayre, J., et al., Nurse visit utilization and blood pressure control: A multi-cohort study in New Zealand. Public Health Nurs, 2022. 39(6): p. 1181–1187.

18. Ye, J., et al., Characteristics and Patterns of Retention in Hypertension Care in Primary Care Settings From the Hypertension Treatment in Nigeria Program. JAMA Network Open, 2022. 5(9): p. e2230025–e2230025.

19. Kim, D., et al., Association between office visit intervals and long-term cardiovascular risk in hypertensive patients. J Clin Hypertens (Greenwich), 2023. 25(8): p. 748–756.

20. Ye, J., et al., Interventions and contextual factors to improve retention in care for patients with hypertension in primary care: Hermeneutic systematic review. Preventive Medicine, 2024: p. 107880.

21. Higashiyama, A., et al., Does self-reported history of hypertension predict cardiovascular death? Comparison with blood pressure measurement in a 19-year prospective study. Journal of Hypertension, 2007. 25(5): p. 959–964.

22. Rothwell, P.M., et al., Prognostic significance of visit-to-visit variability, maximum systolic blood pressure, and episodic hypertension. The Lancet, 2010. 375(9718): p. 895–905.

23. Ye, J., et al., Community-Based Participatory Research and System Dynamics Modeling for Improving Retention in Hypertension Care. JAMA Network Open, 2024. 7(8): p. e2430213–e2430213.

24. Kawasoe, M., et al., Development of a risk prediction score for hypertension incidence using Japanese health checkup data. Hypertension Research, 2022. 45(4): p. 730–740.

25. Deng, X., et al., Development and validation of a nomogram to better predict hypertension based on a 10-year retrospective cohort study in China. Elife, 2021. 10.

26. Feng, Y., et al., Personalized prediction of incident hospitalization for cardiovascular disease in patients with hypertension using machine learning. BMC Med Res Methodol, 2022. 22(1): p. 325.

27. Wang, C.C., T.W. Chu, and J.R. Jang, Next-visit prediction and prevention of hypertension using large-scale routine health checkup data. PLoS One, 2024. 19(11): p. e0313658.

28. Zhang, S., et al., Machine Learning-Based Mortality Prediction in Critically Ill Patients with Hypertension: Comparative Analysis, Fairness, and Interpretability. medRxiv, 2025: p. 2025.04. 05.25325307.

29. Ye, J., Predicting mortality in critically ill patients with hypertension using machine learning and deep learning models. Frontiers in Cardiovascular Medicine, 2025. 12: p. 1568907.

30. Xiang, C., et al., Machine learning-based prediction of disability risk in geriatric patients with hypertension for different time intervals. Archives of Gerontology and Geriatrics, 2023. 105: p. 104835.

31. Mroz, T., et al., Predicting hypertension control using machine learning. PLoS One, 2024. 19(3): p. e0299932.

32. Yun, D., et al., Real-time dual prediction of intradialytic hypotension and hypertension using an explainable deep learning model. Sci Rep, 2023. 13(1): p. 18054.

33. Datta, S., et al., Predicting hypertension onset from longitudinal electronic health records with deep learning. JAMIA Open, 2022. 5(4).

34. Du, J., et al., Developing a hypertension visualization risk prediction system utilizing machine learning and health check-up data. Sci Rep, 2023. 13(1): p. 18953.

35. Kario, K., et al., Hypertension Is Predicted by Both Large and Small Artery Disease. Hypertension, 2019. 73(1): p. 75–83.

36. Gallagher, J., et al., B-Type Natriuretic Peptide and Ventricular Dysfunction in the Prediction of Cardiovascular Events and Death in Hypertension. American Journal of Hypertension, 2017. 31(2): p. 228–234.

37. Chowdhury, E.K., et al., Visit-to-visit (long-term) and ambulatory (short-term) blood pressure variability to predict mortality in an elderly hypertensive population. Journal of Hypertension, 2018. 36(5): p. 1059–1067.

38. Ye, J., Transforming and facilitating health care delivery through social networking platforms: evidences and implications from WeChat. JAMIA open, 2024. 7(2): p. ooae047.

39. Ye, J. and Z. Ren, Examining the impact of sex differences and the COVID-19 pandemic on health and health care: findings from a national cross-sectional study. JAMIA Open, 2022.

40. Ye, J., et al., DeepSeek in Healthcare: A Survey of Capabilities, Risks, and Clinical Applications of Open-Source Large Language Models. arXiv preprint arXiv:2506.01257, 2025.

41. Ye, J., et al., Development and Application of Natural Language Processing on Unstructured Data in Hypertension: A Scoping Review. medRxiv, 2024: p. 2024.02. 27.24303468.

42. Ye, J. and S. Bronstein, Using shared clinical decision support to reduce adverse drug events and improve patient safety. Frontiers in Digital Health, 2025. 7: p. 1703141.

43. Sanuade, O.A., et al., Fixed-dose combination therapy-based protocol compared with free pill combination protocol: Results of a cluster randomized trial. The Journal of Clinical Hypertension, 2023. 25(2): p. 127–136.

44. Ye, J. and Q. Ma. The effects and patterns among mobile health, social determinants, and physical activity: a nationally representative cross-sectional study. in AMIA Annual Symposium Proceedings. 2021. American Medical Informatics Association.

45. Ye, J., et al., The role of artificial intelligence in the application of the integrated electronic health records and patient-generated health data. medRxiv, 2024: p. 2024.05. 01.24306690.

46. Ye, J., et al., Identifying Contextual Factors and Strategies for Practice Facilitation in Primary Care Quality Improvement Using an Informatics-Driven Model: Framework Development and Mixed Methods Case Study. JMIR Human Factors, 2022. 9(2): p. e32174.

47. Conn, V.S., et al., Interventions to improve medication adherence in hypertensive patients: systematic review and meta-analysis. Current hypertension reports, 2015. 17(12): p. 94.

48. Márquez Contreras, E., et al., Specific hypertension smartphone application to improve medication adherence in hypertension: a cluster-randomized trial. Current Medical Research and Opinion, 2019. 35(1): p. 167–173.

49. Zhang, Z., et al., Echo-Vision-FM: a pre-training and fine-tuning framework for echocardiogram video vision foundation model. Nature Communications, 2025.

50. Ye, J., et al., Multimodal data hybrid fusion and natural language processing for clinical prediction models. AMIA Summits on Translational Science Proceedings, 2024. 2024: p. 191.

51. Zhang, W., et al., Organ-aware Multi-scale Medical Image Segmentation Using Text Prompt Engineering. arXiv preprint arXiv:2503.13806, 2025.

52. Ye, J., et al., Implementation outcomes from the Hypertension Treatment in Nigeria program: results from a type 2 hybrid interrupted time series trial. Implementation Science, 2025.

